# FDA oversight of NSIGHT genomic research: the need for an integrated systems approach to regulation

**DOI:** 10.1101/19001370

**Authors:** Laura V. Milko, Flavia Chen, Kee Chan, Amy M. Brower, Pankaj B. Agrawal, Alan H. Beggs, Jonathan S. Berg, Steven E. Brenner, Ingrid A. Holm, Barbara A. Koenig, Richard B. Parad, Cynthia M. Powell, Stephen F. Kingsmore

## Abstract

The National Institutes of Health (NIH) funded the **N**ewborn **S**equencing **I**n **G**enomic medicine and public **H**eal**T**h (NSIGHT) Consortium to investigate the implications, challenges and opportunities associated with the possible use of genomic sequence information in the newborn period. Following announcement of the NSIGHT awardees in 2013, the Food and Drug Administration (FDA) contacted investigators and requested that pre-submissions to investigational device exemptions (IDE) be submitted for the use of genomic sequencing under Title 21 of the Code of Federal Regulations (21 CFR) part 812. IDE regulation permits clinical investigation of medical devices that have not been approved by the FDA. To our knowledge, this marked the first time the FDA determined that NIH-funded clinical genomic research projects are subject to IDE regulation. Here we review the history of and rationale behind FDA oversight of clinical research and the NSIGHT Consortium’s experiences in navigating the IDE process. Overall, NSIGHT investigators found that FDA’s application of existing IDE regulations and medical device definitions aligned imprecisely with the aims of publicly funded exploratory clinical research protocols. IDE risk assessments by the FDA were similar to, but distinct from, protocol risk assessments conducted by local Institutional Review Boards (IRBs), and had the potential to reflect novel oversight of emerging genomic technologies. However, the pre-IDE and IDE process delayed the start of NSIGHT research studies by an average of 10 months, and significantly limited the scope of investigation in two of the four NIH approved projects. Based on the experience of the NSIGHT Consortium, we conclude that policies and practices governing the development and use of novel genomic technologies in clinical research urgently need clarification in order to mitigate potentially conflicting or redundant oversight by IRBs, NIH, FDA, and state authorities.

## INTRODUCTION

In 2013, the Eunice *Kennedy Shriver* National Institute of Child Health and Human Development (NICHD) and the National Human Genome Research Institute (NHGRI) co-founded the Newborn Sequencing In Genomic medicine and public HealTh (NSIGHT) Consortium to better understand the use of genomic testing in the newborn period^1^. Respondents to the request for proposals were asked to address one or more of the following scientific questions: For disorders currently screened for in newborns, how can genomic sequencing replicate or augment known newborn screening results? What knowledge about conditions not currently screened for in newborns could genomic sequencing of newborns provide? And, what additional clinical information could be learned from genomic sequencing relevant to the clinical care of newborns? In addressing these questions, applicants were required to develop a research project with three components: genomic sequencing, clinical research, and ethical, legal, and social implications (ELSI) research. The NICHD and NHGRI ultimately funded the proposals of four academic medical institutions, and here we describe their unique experiences responding to the Food and Drug Administration’s (FDA) unprecedented oversight of NIH-sponsored clinical research projects, and provide insight and recommendations for regulators, researchers, and policy-makers.

### Regulatory Background

The use of genetic and genomic sequencing to guide clinical decision making has rapidly increased, driven by the decreasing cost and turnaround time of next generation sequencing (NGS) technologies, expanding knowledge about the impact of genetic variation on disease and treatment choice, and growing evidence of the benefits of precision medicine to save lives and improve health outcomes. Though the regulation of genetic testing technologies has received attention over the past 25 years, establishing meaningful oversight of genetic and genomic testing has proven complicated. The variable scope, utility, and applications of the data generated through sequencing technologies, as well as interpretive challenges, make protection of research participants and patients at the increasingly blurry boundary between genomic research and clinical care especially complex. Federal agencies tasked with overseeing evolving genomic technologies face the unenviable task of regulating a rapidly moving target^2^. As a result, regulations may lag behind the application of genomic technologies and prove imperfect mechanisms for oversight when applied across diverse use cases.

As recipients of NIH funding, NSIGHT Consortium research is subject to oversight by the Department of Health and Human Services’ (DHHS) Office for Human Research Protection (OHRP) under the Common Rule^3^. DHHS houses the FDA, which derives the authority to regulate medical devices from the 1938 Food, Drug, and Cosmetic Act and subsequent amendments and is tasked with ensuring that tests constituting medical devices are both safe and effective^4^. Within FDA, the Center for Devices and Radiological Health (CDRH) is responsible for regulating the manufacture of *in vitro* diagnostic devices that are sold in the United States, including NGS tests, which are considered a type of medical device under Title 21 of the US Code (USC) section 321. The CDRH issues Investigational Device Exemptions (IDE) to permit the clinical evaluation of investigational devices prior to marketing approval and to concomitantly protect the health and safety of clinical research subjects. However, research studies of diagnostic devices have historically been exempt from the IDE regulations unless the reviewing Institutional Review Board (IRB) has previously determined that a study involves a significant risk device. In addition, much of genomic medicine research involves questions that are focused on the results of genomic sequencing, where the actual “device” used to generate sequencing data is less important than understanding what clinicians and patients do with the information. Therefore, while the full scope of a genomic medicine research project may inevitably involve the generation of sequencing data on a research participant, the intent of the research may not be to validate a test for clinical marketing, but rather to use sequencing technology as a starting point from which to ask broad questions, for example about the ethical, legal, and social implications of genetic findings.

Although the IDE regulation has been in place for over four decades, the FDA’s role in the oversight of genomics research continues to evolve as genomic sequencing is increasingly being applied across a variety of patient populations and healthcare settings. The Secretary’s Advisory Committee on Genetic Testing first called for FDA involvement in the oversight of genetic testing in 2008^5^. Nonetheless, many researchers are not aware of the FDA role in clinical research or the FDA’s categorization of genomic sequencing as a medical device that would require FDA oversight and evaluation to allow for its use in medical research^6^.

In addition, research testing where patient-specific results are reported from a laboratory, and those results will be or could be used “for the diagnosis, prevention, or treatment of any disease or impairment of, or the assessment of the health of, human beings” are subject to Clinical Laboratory Improvement Amendments (CLIA) certification (Title 42 Code of Federal Regulations). CLIA is overseen by the Centers for Medicare and Medicaid Services (CMS), which, like the FDA, is part of the Department of Health and Human Services. The CLIA program in each state is overseen by the respective Department of Health or Public Health. The objective of the CLIA program is to ensure accurate and reliable laboratory test results.

The FDA defines a Laboratory Developed Test (LDT) as an in vitro diagnostic test that is manufactured by and used within a single laboratory (i.e. a laboratory with a single CLIA certificate). LDTs are also sometimes called in-house developed tests, or “home brew” tests. Similar to other in vitro diagnostic tests, LDTs are considered “devices,” and are therefore subject to regulatory oversight by the FDA. When a laboratory develops a test system such as an LDT in-house without receiving FDA clearance or approval, CLIA allows the return of test results following establishment of certain performance specifications relating to analytical validity for the use of that test system in the laboratory’s own environment (see 42 CFR 493.1253(b)(2). This analytical validation is limited, however, to the specific conditions, staff, equipment and patient population of the particular laboratory, so the findings of these laboratory-specific analytical validations are not meaningful outside of the laboratory that did the analysis. Unlike the FDA, whose review of analytical validity is done prior to the marketing of the test system, a laboratory’s analytical validation of LDTs is reviewed by CLIA during its routine biennial survey – after the laboratory has already started testing. Many clinical laboratories utilize the College of American Pathologists (CAP’s) Laboratory Accreditation Program to help meet CLIA requirements. Unlike the FDA regulatory scheme, the CLIA program does not address the clinical validity of any test. Thus, the two agencies’ regulatory schemes are different in focus, scope and purpose, but they are intended to be complementary.

### Oversight of genomic testing used in NIH-sponsored research

In genomic clinical research, the IDE process is focused on the analytical validity of the genomic test and is designed to protect the interests of research participants whose clinical care may be impacted by the results of the genomic test. Research projects that do not return genomic test results to research participants or their physicians are exempt. Studies that use a second “medically established” procedure to confirm the results of the genomic test may also be exempt, but only if they are being used to confirm a variant for which the diagnostic interpretation is incontrovertible^4^. For non-exempt studies, FDA has two classifications of risk: non-significant risk (NSR) and significant risk (SR). The risk determination process focuses on the relative risks posed to the study participants, and the investigator conducting the research makes an assessment of risk that is communicated in an IRB protocol submitted to their IRB. The IRB decides if the study is exempt, NSR, or SR, and the reason for the classification is described in detail. Studies that are determined to be SR require the investigator to submit an IDE application to the FDA before beginning their project, and the IDE application must be approved by the FDA prior to the enrollment of participants.

### FDA Involvement with the NSIGHT consortium

Shortly after the NIH announcement of the NSIGHT awards in late 2013, the Principal Investigators (PI) of the four NSIGHT research groups were informed that they were required to submit a 510(k) Pre-Submission to an IDE application (“Pre-Sub”) for premarket review prior to review by the local IBR, despite a contradictory FDA statement that a Pre-Sub was entirely voluntary on the part of the applicant^7,8^. Subsequently, each of the research groups embarked on an interactive and arduous review process with FDA before beginning enrollment. This entailed correspondence at great length via email, letter, telephone and videoconference with FDA representatives about the submission of the Pre-Sub and, in the case of the NC NEXUS study at the University of North Carolina, Chapel Hill, led to ongoing FDA oversight and interactions after the full IDE submission was approved (Fig. 1). The following sections detail the unique “devices” and the concomitantly unique experiences of the four studies while navigating the unfamiliar and unexpected regulatory oversight of the FDA.

**Figure 1:**
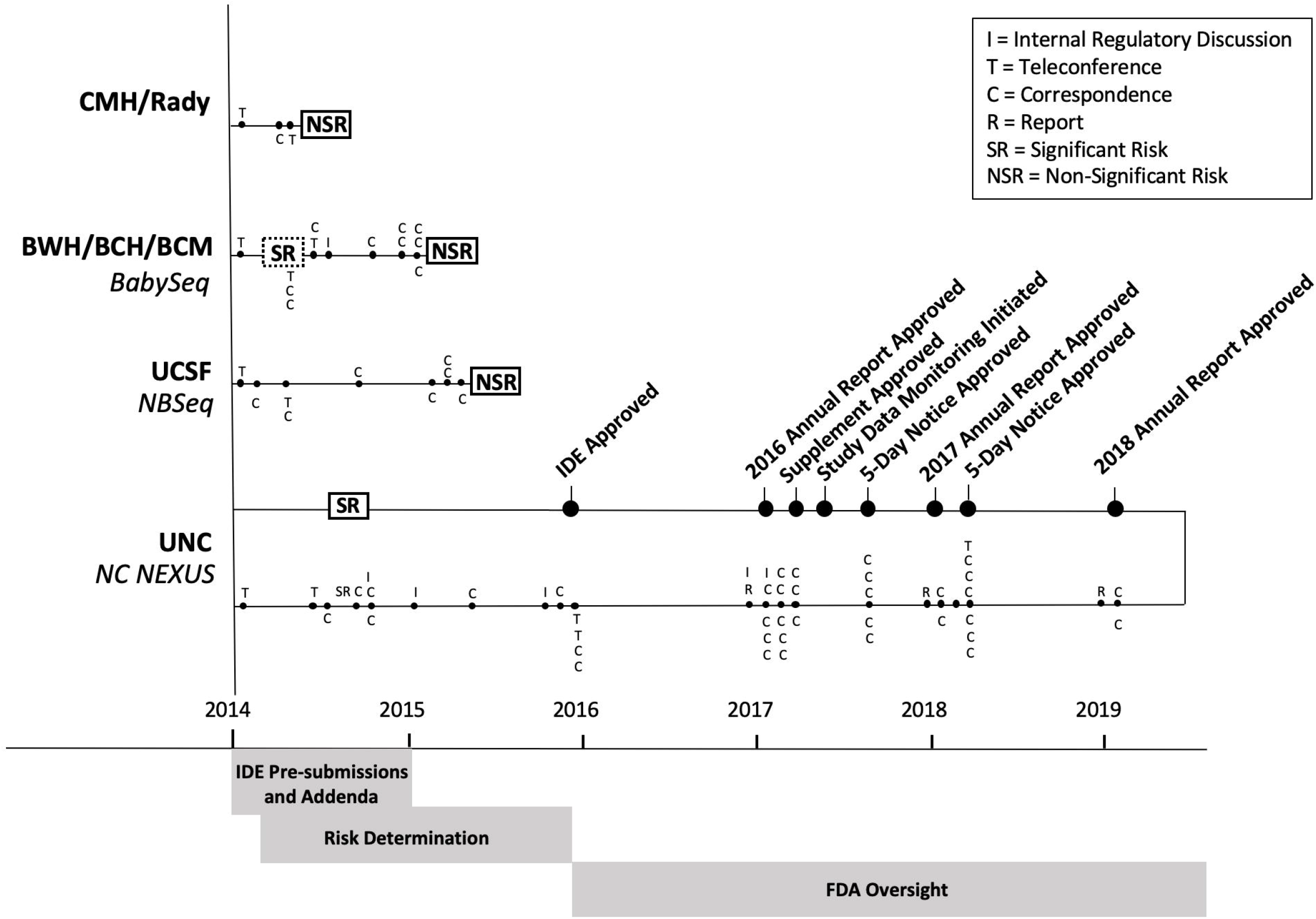
Comparative timelines of FDA interactions and decision making with the 3 sites (Rady, BWH/BCH/BCM and UCSF) that were determined to be Non-Significant Risk (NSR) and with the remaining site (UNC) in which Significant Risk (SR) was determined. The phases of FDA - NSIGHT interaction were: IDE Pre-submission, in which all sites participated and submission of Pre-sub addenda, if needed by FDA; Risk determination; and ongoing FDA oversight for the NC NEXUS study (UNC). FDA-initiated activities are shown below the axis for each site, where as NSIGHT site-initiated activities are indicated above the axis for each site.

## METHODS

### Experiences of NSIGHT investigators with FDA

#### Children’s Mercy Hospital, Kansas City, MO/Rady Children’s Institute for Genomic Medicine, San Diego, CA

Children’s Mercy Hospital (CMH) submitted a Pre-Sub in March 2014 for the first study iteration (NSIGHT1), a partially blinded randomized control trial (RCT) of rapid whole genome sequencing (WGS) plus standard tests versus standard tests alone.^9-14^ The participants were families with an infant aged <4 months in a regional neonatal intensive care unit (NICU) or pediatric intensive care unit (PICU) with an illness of suspected genetic etiology. The primary end-point was the rate of etiologic diagnosis within 28 days of test order.

The device comprised innovative methods for making a rapid diagnosis of a simple genetic disease, including deep phenotyping, rapid whole genome sequencing (rWGS), rapid primary, secondary and tertiary sequence analysis, diagnostic interpretation, confirmatory testing, reporting results, and provision of inpatient precision medicine guidance. Most of the device elements had previously been inspected by CLIA and CAP and been found compliant, including rWGS performed at the CMH CLIA laboratory under local IRB research protocols.^10^ Unpublished data regarding the diagnostic and clinical utility of rWGS was included in the Pre-Sub.^12,13^

The FDA’s initial review of the CMH study was received on April 28, 2014, and requested additional information regarding maximum blood draws in neonates, and regarding verbal provisional return of diagnostic results prior to confirmation by an orthogonal method. The study clarified that the latter was limited to NICU and PICU infants with life-threatening conditions in whom the molecular diagnosis was associated with a specific treatment that would have a high likelihood of improving outcomes. The potential harm of delayed reporting and institution of specific treatment (1-2 weeks for confirmatory testing) was asserted to be greater than that of a false positive result in this patient population, and had been substantiated by an unpublished case example where returning a verbal, provisional diagnosis had likely saved that infant’s life. Of note, all diagnoses were confirmed prior to final reporting. The FDA determined on May 8, 2014 that the study was NSR and did not require an IDE.

In June 2016, upon transfer of the award from CMH to the Rady Children’s Institute for Genomic Medicine (RCIGM), a second RCT (NSIGHT2), comparing the clinical utility of singleton and trio rWGS with that of singleton and trio rapid whole exome sequencing (WES) was initiated. A number of the device elements differed between CMH and Rady. The RCIGM laboratory had previously been inspected by CLIA and CAP and been found compliant, including rWGS. The FDA was contacted regarding whether another Pre-Sub was required for the new enrollment site and clinical laboratory, and about interim changes to the software and hardware components of the device that improved analytic performance.^14^ The FDA determined that another Pre-Sub was not necessary since the risk determination had not changed materially.

#### “NBSeq” study at the University of California, San Francisco

The NBSeq project, as described in the Pre-Sub submitted in January 2014, was designed to evaluate the potential application of WES in newborn screening (NBS) using 1) deidentified archived dried blood spots (DBS) to ascertain metabolic disorders currently screened for in the state of California, and 2) identified archived DBS, obtained with parental consent, from individuals with an immunodeficiency disorder not identified by current NBS. The deidentified DBS research was not subject to FDA oversight, as no results would be returned. In the immunodeficiency cohort, the NBSeq project proposed to use WES to identify variants potentially associated with the child’s immune disorder. Parents provided informed consent for testing. The NBSeq investigators also initially planned to explore certain pharmacogenomic (PGX) variants as “secondary” findings, and offer parents the option to receive those results. The team also sought to capture prospectively the decision process, including parental reactions to results and opinions regarding the value to their family of WES.

The device was considered to be DNA extraction and exome sequencing using the Illumina HiSeq 2500 system, with clinical confirmation prior to return of results. Interesting PGX variants identified via WES would be confirmed in a CLIA-certified commercial laboratory using a cost-effective LDT on a clinically routine genotyping platform and only validated results would be shared with parents who had consented to receive results.

Initially, the FDA indicated that the NBSeq project would be a SR study, despite the routine use of the intended confirmatory PGX test in clinical practice to guide medical decisions, because the commercial CLIA-certified method neither used Sanger sequencing nor furnished specific validation data required by the FDA for research studies. An SR determination would have required the study to adopt a much more expensive PGX genotyping method to perhaps render it acceptable to the FDA, to submit a full IDE application, or to change the protocol, eliminating the plan to return PGX results as a secondary finding. In April 2015, the NBSeq investigators decided to eliminate the PGX arm of the research and the FDA subsequently deemed that the study was exempt from IDE regulations.

#### “BabySeq” - Brigham Women’s Hospital and Boston Children’s Hospital, Boston, MA and Baylor College of Medicine, Houston, TX

In their February 2014 Pre-Sub, the BabySeq project described their “Exome Sequencing Test” as falling within the remit of currently described CLIA LDTs, and therefore exempt from regulation as a medical device. The BabySeq project was an RCT designed to investigate the potential impact of genomic information on future care and on the ELSI of returning genomic information, including carrier status, to families and health care providers. A cohort of healthy newborns and a cohort of sick newborns were randomized to a control arm (conventional NBS results and a detailed family history) or an experimental arm (genomic sequencing in addition to conventional NBS and a detailed family history). Surveys of parents and physicians assessed attitudes and preferences, health care utilization, health behaviors and intentions, decisional satisfaction, and psychosocial impact on the family.

The device description included routine clinical specimen collection and DNA extraction, genomic sequencing on an Illumina MiSeq Sequencer, sequence variant annotation and interpretation utilizing industry standard software and following the American College of Medical Genetics and Genomics (ACMG) and Association of Molecular Pathologists (AMP) professional guidelines.^15^ Orthogonal confirmation of positive results would utilize independent sequencing on an Ion Torrent Proton Sequencer or by traditional Sanger methods, and Droplet Digital PCR (ddPCR) confirmation of potential copy number variants would be performed in CLIA/CAP compliant clinical diagnostic laboratories. Board certified genetic counselors in conjunction with a study physician would return results to participants.

In May 2014, the FDA responded that the “Test”, though developed and validated in the same way as other LDTs, did not qualify as NSR because neither the NGS nor the ddPCR orthogonal confirmation had prior FDA approval and due to the undefined nature of the thousands of genes being queried, it was not possible to anticipate in advance the medical conditions and treatment decisions that would be encountered.

In December 2014, after extensive deliberation, consultation with hospital regulatory experts, and protocol approval by local IRBs, the BabySeq study team submitted an addendum to the initial Pre-Sub that included several key changes and clarifications: 1) all variants found by NGS would be confirmed by Sanger sequencing; 2) only genes with definitive or strong evidence to cause pediatric onset conditions (but not adult onset conditions as originally proposed) would be adjudicated and included in reports; and 3) only variants meeting ACMG/AMP criteria for pathogenicity would be reported. In addition, due to cost constraints, with the added requirement for Sanger sequencing confirmation, reporting of blood typing results was dropped from the protocol. The FDA rendered a final determination that the BabySeq program represented an NSR device study on February 23, 2015 based on the three previous factors and the clarification that result disclosure would be conducted via appropriate physician and genetic counselor interactions.

#### “NC NEXUS” - University of North Carolina, Chapel Hill

The NC NEXUS project submitted the Pre-Sub in January 2014 describing the study as an embedded two-arm, parallel group, RCT of two cohorts of children: one cohort with a condition recently diagnosed through standard NBS and one cohort of newborns identified during a healthy pregnancy^16^. Parents of both cohorts were consented to learn the results of an “NGS-NBS” panel of medically actionable conditions comparable to those detected by current NBS screening^17^, and also to be randomized to have the ability to request additional genomic findings (“Decision Arm”) or not (“Control Arm). Recruitment of both healthy and diagnosed cohorts facilitated the study of parental decision-making and a novel evaluation of the potential ability of NGS-NBS to enhance and augment current NBS.

Initially the NC NEXUS investigators defined the device as including the exome sequencing methods, bioinformatics pipeline, and variant calling software, as well as confirmation of variants using Sanger sequencing in a CLIA-certified clinical laboratory and reporting by a board-certified molecular pathologist. However, several weeks after receiving the Pre-Sub, FDA reviewers requested an addendum to the Pre-Sub describing the ELSI arm of the device, including specifics about the electronic decision aid being developed to assist parents in making decisions about exome sequencing in their child^18^, the populations to be studied, the genes to be analyzed, and the types of variants that would be reported.

After extensive correspondence between UNC investigators and FDA reviewers, FDA indicated that a primary concern was that the “Decision Arm” would have the option of learning additional results from the exome sequencing data that would include pathogenic variants in genes associated with adult onset medically actionable conditions. Although the NC NEXUS study noted that board certified generic counselors and clinical geneticists would conduct visits for informed consent and return of results, FDA’s concerns were not completely offset.

The FDA expressed additional concerns, including how to define a “gold standard” negative result and whether extensive validation of the device had been conducted. Since one of the aims of the study was to evaluate the performance of exome sequencing in a screening context, no such validation had been performed. In addition, NC NEXUS investigators noted that the main scientific goals were to study parental decision-making about genomic findings in newborns and children, and not to validate a test that would subsequently be offered as a clinical service or marketed and commercialized. Eventually, the FDA was convinced that developing an appropriately validated negative result would require a much larger study and was outside the scope of the NC NEXUS study.

On August 28, 2014, the FDA concluded that the NC NEXUS study was SR and required a full IDE submission prior to beginning recruitment and enrollment. At this time, the NC NEXUS PIs chose to submit a full IDE rather than significantly alter their research proposal. Development of all the study materials that were required by the IDE, including protocol workflows, recruitment materials, consent forms, reporting forms, on-line decision aid tools, and the list of genes that would be included in the study, were part of the project and allowed to proceed; however, recruitment and enrollment of study participants was stalled pending FDA approval of the IDE, which was finally received on December 22, 2015.

## RESULTS

Protecting human participants who are enrolled in research studies involving significant risk medical devices is a critically important role of the FDA. However, determining whether the level of risk warrants an IDE application to FDA and, possibly, future FDA oversight is the responsibility of the project investigator’s IRB. Despite a clear statutory requirement that the investigator’s IRB should make the risk determination, the FDA pre-emptively requested Pre-Subs from the NSIGHT investigators and made risk determinations, which for some projects necessitated profound changes to their funded study proposals, before the protocols were submitted to the local IRBs. Because of this atypical sequence of risk determination and a lack of specific guidelines regarding the risk determination criteria, the NSIGHT consortium engaged in a lengthy and arduous process of understanding unfamiliar FDA procedures and attending to compliance. NSIGHT researchers ultimately identified several common elements within our unique study designs which we believe served as the basis for the FDA’s risk determinations.

### Elements of Risk Determination

The FDA’s accumulated experience with IDE submissions for other medical devices informed the development of an internal rubric that is applicable across different studies; however, the specific details of the criteria for risk determination for genomic sequencing studies have not yet been articulated to our knowledge. Though the four studies had unique designs and clinical contexts, all had proposed to investigate the application and performance of NGS approaches in newborns and children (per the mission of the consortium), and all had originally planned to return results to study participants in order to study the downstream implications of doing so. Through the collective experience of the consortium, we identified four factors we believe contributed the greatest impact in the FDA’s risk determination: 1) method of orthogonal confirmation; 2) method of return of results; 3) population to whom results are returned; and 4) specific types of results being returned.

Confirmatory testing methods varied for each of the studies because the NSIGHT consortium was established for the purpose of investigating the application and performance of NGS (exome or genome sequencing) in the diagnosis and screening of newborns and children. Historically, Sanger sequencing has been the “gold standard” and it was made clear that other methods, such as confirmation of potential copy number variants using ddPCR as initially proposed by the BabySeq project, were deemed unacceptable without prior extensive and cost-prohibitive validation. Subsequent studies have shown that Sanger sequencing is subject to the same amplification and repeat-based artifacts as NGS and is not necessary for confirmation of all NGS results.^19–21^

The FDA also scrutinized the method by which results would be returned to the participants. Studies that utilized a genetic counselor, clinical geneticist, and/or study physician in the return of results were deemed of lower risk than studies that provided results without counseling. However, the FDA communicated that genetic counseling, while considered to be a mitigating factor, was not sufficient in and of itself to make a determination of NSR. Research studies in which results are returned to participants by other methods are construed as posing increased risk, presumably due to the chance for misunderstanding.

Though the mandate of the NSIGHT consortium was to investigate the applications and efficacy of genomic sequencing in infants and children, the return of results in those populations appeared to play a major role in the risk determination. In particular, the FDA considered whether the results were being returned in a population already affected with certain health conditions (eg. seriously ill NICU patients) or those who were healthy. In the FDA’s view, return of results in the healthy population conferred a higher level of risk that could not be mitigated by the precautions that are in place as part of routine medical care. The fact that the research participants were children also appeared to increase the overall level of risk.

The FDA was also interested in the inherent risks to returning different types of diagnostic and public health screening results. The Rady Children’s Institute/CMH study was designed to investigate the efficacy of analyzing and returning genomic diagnostic results, which had been shown to have demonstrable benefits and an overall low risk of harm, to critically ill newborns in the NICU and PICU^11,22^. Conversely, the BabySeq, NBSeq, and NC NEXUS studies proposed to examine the application and efficacy of genome-scale sequencing in the context of returning public health screening results to the parents of healthy newborns. Both the BabySeq and NC NEXUS studies originally proposed to return results for conditions with likely childhood onset as well as adult onset. These results were categorized in terms of the gene-disease pairs that would be analyzed for possible return of results in the study, as well as the interpretive classifications of variants that would be returned (e.g. pathogenic, likely pathogenic, variant of uncertain significance). Each NSIGHT group defined their return of results protocols differently, depending on the aims of the study, which in turn influenced the risk determination.

Considering these factors together, we can draw some conclusions about the FDA’s risk determination process for the NSIGHT Consortium studies. The FDA appeared to consider returning diagnostic results to parents of infants or children with an existing health condition to be lower risk than returning secondary findings or primary screening results to parents of asymptomatic newborns. Additionally, returning results to parents of infants or children that are related to childhood onset conditions is considered lower risk than returning results related to adult onset conditions; this is likely due to the FDA’s interpretation of ethical guidance regarding the use of genetic tests in minors.^23,24^ A further layer of complexity in the risk assessment was the intersection between the interpretation of variants (the criteria that would be used and the credentials of the individuals responsible), the classes of variants that would be returned (e.g. pathogenic variants versus variants of uncertain significance) and the clinical context in which those results would be disclosed (diagnostic findings versus screening). The NC NEXUS study, which included the return of results related to adult onset conditions in healthy newborns, was deemed to be SR. The BabySeq study was deemed to be NSR once it altered its protocol and agreed to only report variants associated with pediatric onset conditions and only in genes with “definitive” or “strong” levels of evidence for association with disease.

In the FDA’s interpretation of risk, it seems that the nature of the risk (as defined by the above criteria) is the defining characteristic, and not the likelihood that any harms would actually occur. In the NC NEXUS and BabySeq studies the populations are small and the potential genetic conditions identified are rare. Therefore, there was a very small a priori chance that any given condition would be identified and therefore returned. However, the very low likelihood did not mitigate the risk.

### Impact of Risk Determination

All four sites were significantly impacted by the mandatory Pre-Sub process, including delayed submission and review by IRBs, and onset of clinical enrollment that resulted in decreased enrollment and the alteration of originally planned protocols, as a result of the unexpected demand on researchers’ limited resources to interpret and navigate the FDA requirements. Ultimately, NBSeq and BabySeq substantially curtailed their study design and protocols, to the detriment of the entire consortium-wide research directive, in order to avoid the subsequent resource-intensive IDE submission process. The NBSeq study investigators ultimately eliminated the PGX aspect of their study due to a lack of resources to generate an FDA compliant Pre-Sub, to avoid further delay in the conduct of the research, and the proportionally small number of families expected to be studied as part of the PGX project.

The BabySeq study made several substantive modifications to their originally proposed and funded protocol. Due to FDA requirement for Sanger confirmations, it became financially and logistically unfeasible to return non-medically critical findings, such as minor blood group antigens and PGX findings, so the region of interest and scope of the study was effectively constrained. The study also proceeded without the initial aim to confirm smaller copy number variants by ddPCR. The restriction on returning results only for pediatric onset or actionable conditions led to a dilemma later on when an infant was found to harbor a maternally inherited adult-onset cancer risk allele, necessitating an urgent protocol amendment and FDA notification.^25^ Furthermore, the lengthy regulatory delay, with the loss of over 12 months of funded time blocked to recruitment, contributed to the diminished enrollment of only 316 of the proposed 480 newborns prior to the study end date.^26^

The SR determination levied against the NC NEXUS study, and the subsequent decision by the study PIs to submit a full IDE application rather than substantially alter their research aims and protocol, delayed the NC NEXUS study by more than 12 months, negatively impacted recruitment and enrollment of participants and added substantial unanticipated costs. The analysis was restricted to small variants that could be confirmed by Sanger sequencing, rather than the original plan of analyzing both small variants and predicted copy number variants based on depth of coverage analysis, due to the complications of determining and validating a confirmatory test strategy acceptable to the FDA. Of note, this meant that the diagnostic yield of the research exome was intentionally restricted by the investigators due to the challenges of FDA oversight. Following approval of the NC NEXUS IDE more than 2 years after the initial notice of award, there were numerous unanticipated administrative requirements that impacted the UNC study in major and minor ways, such as: 1) changes to the original patient consent forms; 2) changes to the original plans for how “negative” screening reports would appear; 3) hiring an independent data monitor consultant to review study data on a regular basis; 4) regular reporting through IDE annual reports; and 5) study protocol addenda that had to be submitted to the FDA prior to implementation (either 5-day notices of minor procedural changes or more detailed supplemental applications for changes such as modifications to the exome capture protocol or to have the ability to enroll parents of children with variants of unknown significance in order to perform family segregation studies).

In contrast, the NSR determination for the Rady/CMH study was considered beneficial by the PIs in several ways. Firstly, the process of risk determination by the FDA was helpful, providing a different, more analytically rigorous perspective that helped inform the subsequent IRB protocol. Likewise, the CMH IRB found the NSR determination by the FDA to be helpful in their review of what was considered a highly innovative protocol, particularly from an ELSI standpoint. Finally, when the study moved from CMH to Rady, with testing at a different laboratory and with different “device” components, notification by the FDA that the “device” did not require an additional Pre-IDE submission was helpful in accelerating review by the Rady IRB.

## DISCUSSION

Clinical genomic research is funded and regulated by many federal, state and local (institutional) agencies. As evidenced by the NSIGHT program, the boundaries of oversight by these agencies are not always apparent to researchers or sister agencies, and may conflict with one another. We found that the basis for risk assessments and prioritization of areas of concern with regard to novel genomic technologies varied considerably between the NIH, FDA, CMS (CLIA), and local IRBs. Different stakeholders worked in tandem, but their efforts were isolated from each other and this uncoordinated regulation of the NSIGHT consortium resulted in an average delay of approximately 12 months, additional cost and drain on resources, and change in Specific Aims in two of the four studies. Given the typical 4-5 year duration of NIH-funded research, such delays curtailed enrollment, which weakened the statistical power of studies and decreased the likelihood that Specific Aims were achieved. If delays had resulted in corresponding risk reduction, they would have been viewed as justifiable, but this was not the case for any of the four studies.

While FDA reviewers with appropriate backgrounds may potentially provide deeper expertise with cutting-edge genomic technologies than local IRB reviewers, the transition from evaluating analytic validation to evaluating clinical genomic analyses proved to be a steep learning curve for several FDA reviewers, with clear evidence that the interaction with the NSIGHT groups provided a great deal of insight into this field. Unfortunately, turnover among the FDA review teams resulted in the need for frequent re-education by the NSIGHT investigators. This experience clearly demonstrates that clinical researchers, funding agencies, the general public, local IRBs, and government oversight agencies (i.e. FDA, CMS and CDC) would benefit from a more integrated systems approach to oversight (Fig. 2)^27,28^.

**Figure 2:**
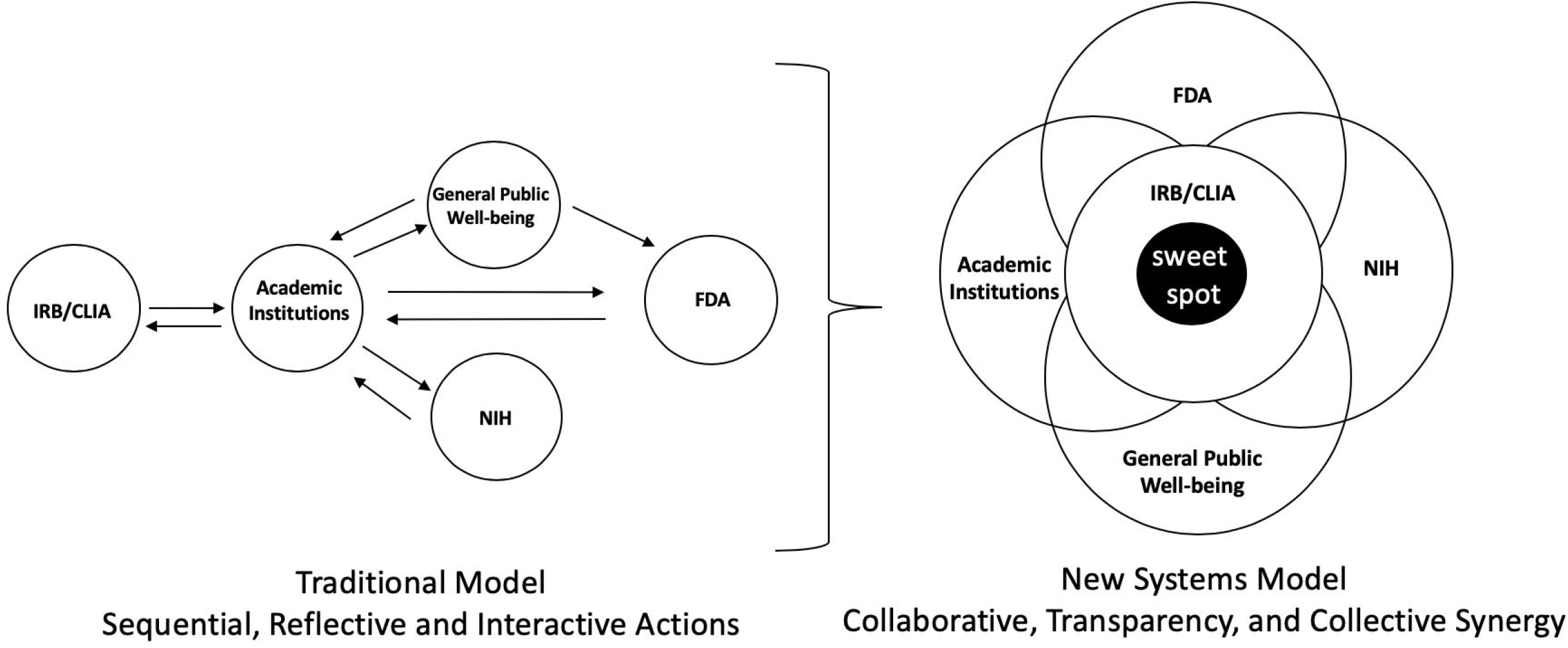
Transition from the traditional framework to a systems approach for oversight of clinical research. A. Traditional oversight involves sequential, uncoordinated actions by various agencies. Principal investigators (PIs) seek general public well-being by applying for funding from NIH to perform clinical genomic research. Public announcements of awards and research projects resulted in FDA awareness of NIH-funded research and FDA expression of concerns of risk assessment, benefits and harms, and clinical utility. Local IRBs and state CLIA requirements provided independent oversight. B. A proposed model of collaborative, transparent, collective research oversight in which the six entities involved (PIs, NIH, FDA, local IRBs, CLIA and the General Public) communicate effectively with regard to benefits and harms. The solid centered blue is the “sweet spot”.

Most NIH research involves ongoing methods development and refinement during the period of award. This is particularly true of studies involving NGS platforms. During the period of NSIGHT awards the underpinning technology platforms have evolved considerably. For example, sequencers manufactured by Illumina Inc., an industry standard, have evolved from the HiSeq 2000 to the HiSeq 2500, then HiSeq 4000 and finally the NovaSeq 6000, with different quality, throughput and turnaround time. Similar improvements have occurred in the analysis software used to call variants (including greater ability to identify structural and copy number variations and even triplet repeat expansions), and the data sources that allow molecular geneticists to determine which variants are clinically relevant. A significant problem with the FDA assessments that occurred at the beginning of studies is that they occurred as a “one off” for a project, and were not well designed to react to such advances in knowledge or changes in technology during the typical 5-year duration of a project. Furthermore, FDA practices also evolved during the period, as evidenced by recent reversals of FDA decisions regarding direct-to-consumer genetic testing^29^. It would seem that IRBs and CMS (through CLIA) oversight mechanisms are better attuned to the tempo of change in biomedical research, with their typical requirement for annual re-approval of protocols and change control mechanisms, respectively.

Based on the collective experiences of the NSIGHT investigators, we propose an integrated approach to the oversight of clinical research using genomic testing, with the short-term goal of protecting research participants’ health while maintaining the long-term goal of improving population health through scientific and clinical discoveries that begins with NIH. We propose that NIH Institutes (and other federal funding agencies) discuss large, new programs with oversight agencies before their implementation, as currently occurs within NIH Institute Councils. Resultant feedback from federal stakeholder agencies, such as CMS and FDA, would assist in drafting and refinement of new program requests for applications (RFA) for clinical genomic research, and with the goal of identifying and addressing boundary issues that were likely to create conflicts or impede programmatic goals or timelines. In addition to the development of RFAs, the relevant oversight agencies would disseminate guidance to investigators considering responses to RFAs or work with NIH staff to include guidance in the RFAs. Such guidance would outline the role that the FDA might play in each particular RFA and assist in determinations such as whether the IDE application pre-approval submission process is mandatory, highly recommended (but optional), or not necessary. In addition, to determine whether FDA oversight is needed for NIH funded genomics clinical research, policies should be established that address more clearly the level of involvement and follow-up required. This could build upon the NSIGHT experience with regard to the level of oversight based on the population under investigation (e.g. adult versus pediatric, healthy versus acutely ill) and the maturity of the genomic technologies, or devices, to be used. Research teams could then use this information as they design studies in response to RFAs. This approach would enable research teams to proactively address identified issues and include funds, expertise and time in proposals to address FDA oversight. During peer review, funding decision making, and upon issuance of an award, the reviewers, the research team and the relevant funding and oversight agencies would then have a clear understanding of the involvement of FDA, and can proceed with IDE pre-submission application and/or IRB protocol submission to their IRB. This approach will also avoid the need to change research protocols after favorable peer review and awarding of funds.

Genomic technologies, including both hardware and software, evolve extremely rapidly, and the potential benefits and harms of exploratory use of novel technologies in clinical research can be extremely difficult to estimate in the absence of data. Therefore, expert consensus guidance is helpful in such determinations but is often limited to extrapolation based on previous experience. When genomic findings are confirmed by an effective orthogonal method, technical false positives can be minimized. Interpretation of the clinical significance of the finding by a qualified practitioner (molecular genetics or cytogenetics) lessens the chance of a clinical false positive. Disclosure to participants via a method that would be deemed standard of care for that type of result should reduce concerns about misunderstanding. Thus, if each of these three features are present, the original technology used to derive the result should be of lesser importance with respect to “positive” findings that are being disclosed to participants. On the other hand, a significant challenge with new technologies is the ability to detect relevant genetic variants compared to other alternatives. Validating a “negative” result becomes extremely difficult, if not impossible in the case of genome-scale sequencing, when the new technology has no equal comparator. For research projects that are not focused on analytical validation, therefore, it may be extremely challenging to provide the types of validation data that would normally be expected by a regulatory agency such as the FDA. One solution to this problem would be to create a new type of targeted NIH research program specifically designed to generate scoping data for novel genomic technologies with regard to, for example, analytic performance or public perception of risk. Results of such evaluations would provide a highly valuable reference framework for investigators and regulatory agencies that would greatly improve the efficiency of review and implementation.

In conclusion, the FDA has authority under 21 CFR 812.20 to regulate NIH-funded clinical research. The FDA exercised this authority in the NHGRI/NICHD NSIGHT program, utilizing the IDE mechanism to evaluate the relative benefits and harms of the human participants research protocols developed by the four grantees, but the RFAs did not address these regulatory aspects. We propose a systems approach that is a coordination between NIH and FDA when planning funding opportunities with the goal of understanding whether the proposed effort will likely require interaction with the FDA. Awareness of the scope of a potential FDA interaction would allow for appropriate budgeting and adjustment of timelines to account for the clearance of potential FDA hurdles. Communication with FDA will enable NIH to provide clearer guidelines to investigators as they design their study, while coordination with NIH will enable FDA to establish a consistent process of review. Provision by the FDA of IDE protocol templates specific to projects that include disclosure of genomic sequencing results, would greatly facilitate researchers’ ability to comply with any regulatory oversight. Here also exists a need for clarification of policies with regard to the potentially conflicting, overlapping oversight of clinical research by local IRBs, NIH, FDA, and state CLIA programs, particularly in light of shifting priorities. The NSIGHT experience highlights the importance of NIH leadership and identifies areas of collaboration and planning that would facilitate successful genomic medicine projects.

By establishing a value-added partnership between academic institutions, NIH, FDA and the general public, we can identify our shared responsibilities and goals. With a shared responsibility and collective systems approach to oversight of genomic research in the future, we may be able to accelerate the adoption of genomics for patient care and public health.

## Data Availability

No data is referred to in the manuscript.

## ACKNOWLEDGEMENTS

The NSIGHT consortium was funded by National Institute of Health grants U19HD077693, U19HD077632, U19HD077627, and U19HD077671. The Newborn Screening Translational Research Network is funded through a contract from the *Eunice Kennedy Shriver* National Institute of Child Health and Human Development (HHSN275201800005C). The authors wish to acknowledge the contributions made to NSIGHT by current and former NHGRI and NICHD Directors Dr. Eric Green, Dr. Teri Manolio, Dr. Alan Guttmacher, and Dr. Diana Bianchi, Program Officers Dr. Anastasia Wise, Dr. Tiina Urv, and Dr. Melissa Parisi, and Scientific Program Analysts Brenda Iglesias, Ellen Howerton, and Cecelia Tamburro.

## AUTHOR CONTRIBUTIONS

L.V.M., S.K., J.S.B., A.M.B., R.B.P, C.M.P., and F.C. contributed to conceptualizing the study and drafting the initial manuscript. P.B.A., I.A.H., and A.H.B contributed to substantive review and revision of the manuscript; L.V.M., J.S.B. and K.C. contributed to the design of the figures A.H.B. drafted and I.A.H. contributed to the description of the BabySeq site experience. L.V.M., J.S.B. and C.M.P. drafted the description of the NC NEXUS study. F.C., B.K. and S.E.B drafted the description of the NBSeq study. All authors read and approved the final manuscript.

